# Comparing Two Food is Medicine Trials Using the RE-AIM Framework

**DOI:** 10.1101/2025.10.17.25338260

**Authors:** Bridget Simon-Friedt, Rachel G. Tabak, Sylvester Tumusiime, Lindsey Rudov, Sang Gune K. Yoo, Stephanie Mazzucca-Ragan, Allison Primo, Derek Hashimoto, Adam Hively, Lingxiu Dong, Mark D. Huffman, Thomas W. Carton, Jing Li

## Abstract

**Introduction:** Food is Medicine (FIM) interventions, such as medically tailored meals, groceries, and produce prescriptions, are increasingly embedded in healthcare delivery. To inform future policy and practice, the American Heart Association’s *Healthcare by Food* initiative supported pilot studies to test scalable FIM models. This manuscript compares two such pilots, NutriConnect (Washington University in St. Louis) and Makin’ Healthy Groceries (Louisiana Public Health Institute), using the RE-AIM framework to highlight cross-site lessons for design, delivery, and implementation.

**Methods:** We conducted a comparative analysis of two FIM trials. NutriConnect enrolled adults ≥18 years recently discharged from Barnes-Jewish Hospital with diet-sensitive chronic conditions and food insecurity, randomizing participants to digital coupons, home-delivered produce boxes, and usual care. Makin’ Healthy Groceries enrolled adults ≥50 years with uncontrolled hypertension at University Medical Center in New Orleans, randomizing participants to in-store debit vouchers or online grocery credits. Across both studies, we applied RE-AIM domains (Reach, Effectiveness, Adoption, Implementation, Maintenance) to examine participant characteristics, intervention delivery, operational challenges, and contextual facilitators.

**Results:** Reach was constrained by digital barriers in both trials: NutriConnect participants struggled with email coupon redemption, while Makin’ Healthy Groceries’ online arm faced low digital literacy and payment concerns. Effectiveness data were collected through validated dietary questionnaires; full quantitative results will be reported separately. Adoption depended heavily on retail system readiness, NutriConnect benefited from tighter integration with the grocer, Schnucks’, loyalty program, while Makin’ Healthy Groceries encountered gaps in staff training and voucher controls at the participating grocery. Implementation challenges included high staff burden for manual troubleshooting and rapid customization of digital platforms, though both studies demonstrated strong adaptive capacity. Maintenance challenges included high program costs and reliance on sustained funding, yet technical enhancements (e.g., automated coupon systems) showed potential for broader scalability.

**Conclusion:** This comparison highlights the heterogeneity of FIM trial design and delivery, underscoring the importance of aligning interventions with participant behaviors, retail system readiness, and digital accessibility. Successful scale-up will require hybrid models that combine technology with human support, strong cross-sector partnerships, and sustainable reimbursement pathways. Insights from these pilots inform the next generation of equitable FIM implementation strategies.

## INTRODUCTION

Nutrition, food access, and dietary behavior are foundational to health outcomes.^1,2^ The growing field of Food is Medicine (FIM) reflects a systematic effort to embed food-based interventions such as medically tailored meals, groceries, and produce prescriptions within healthcare delivery.^3^ In 2022, the American Heart Association (AHA) launched the *Healthcare by Food* initiative to generate evidence on scalable models that improve nutrition, food access equity, health outcomes, and inform health insurance reimbursement policies (by reducing healthcare costs).^4,5^

Through this initiative, the AHA funded a national cohort of pilot FIM programs across 20 institutions. While prior studies have shown modest effects of FIM interventions on health behaviors,^6-10^ *Healthcare by Food* pilots offer an opportunity to analyze variations in current implementation methods.^11-13^ *NutriConnect* led by Washington University in St Louis,^14^ MO (WashU) and *Makin’ Healthy Groceries* led by Louisiana Public Health Institute, in New Orleans, LA (LPHI) are two pilot studies funded by this initiative. Both pilots sought to address food insecurity, promote fruit and vegetable (F&V) consumption, and improve cardiovascular-related health outcomes; however, the programs differed in execution and implementation.

This manuscript compares these two FIM pilots using the RE-AIM framework^15^ to examine differences in design, delivery strategies, and implementation features with a focus on understanding shared challenges and extracting cross-site lessons. The RE-AIM framework is a valuable tool for analyzing public health programs because it provides a comprehensive structure for understanding both the effectiveness and applicability of interventions. This framework supports investigators in identifying important characteristics of interventions that may be generalizable to future FIM programs. While prior research on FIM programs has focused on effectiveness of the intervention on health behaviors and clinical outcomes, we expand our focus to highlight how implementation, when optimized, may foster sustainable strategies that contribute to long-term health effects that are feasible, scalable, and meet population needs.

## METHODS

Both study teams strategically partnered with established regional retail chains as well as a widely utilized online grocery shopping and delivery platform to embed the FIM intervention into natural grocery shopping behavior in order to enhance program feasibility, scalability, and sustainability.

Despite shared goals of improving nutrition and food security, the two pilot studies differed in terms of the target population, enrollment strategy, delivery infrastructure, and intervention arms. These differences create a valuable opportunity to evaluate heterogeneity in real-world delivery of FIM intervention programs.

### Study Contexts and Grocery Partners

- NutriConnect was implemented in St. Louis, Missouri, in partnership with Schnucks, a regional grocery chain with over 100 locations across Missouri, Illinois, Indiana, and Wisconsin, including approximately 50 in the greater St. Louis metropolitan area.
- Makin’ Healthy Groceries was implemented in New Orleans, Louisiana, in partnership with a regional Gulf Coast grocery chain with 15 stores in the New Orleans metropolitan area.

### Participant Profiles and Recruitment Settings

- NutriConnect recruited adult participants 18 years old and older following hospital discharge from Barnes-Jewish Hospital, a large tertiary/quaternary safety-net academic hospital in St. Louis, Missouri. The study targeted individuals with a high burden of chronic, diet-sensitive diseases (e.g., diabetes, hypertension, hyperlipidemia, or obesity) and social needs (i.e., food or financial insecurity). Potential participants were identified through the hospital’s electronic medical record and recruited by clinical social work and study teams. As part of routine care, health workers screened participants for food security, and these data were included in the medical record. Patients with food insecurity were eligible to be recruited to the study.
- Makin’ Healthy Groceries enrolled participants from University Medical Center (UMCNO), New Orleans, Louisiana. The target population included adults 50 years and older with a diagnosis of uncontrolled hypertension defined by at least two clinically documented blood pressure readings greater than 140/90 mmHg within the prior 6 months. Many participants were long-standing ambulatory patients in the UMCNO safety-net health system. Eligible participants were contacted through electronic health record message and enrolled by study coordinators.

Differences in the target population, recruitment strategy, and participant acuity influenced engagement capacity and intervention acceptability. Table 1 provides an overview of each study.

**Table 1.**
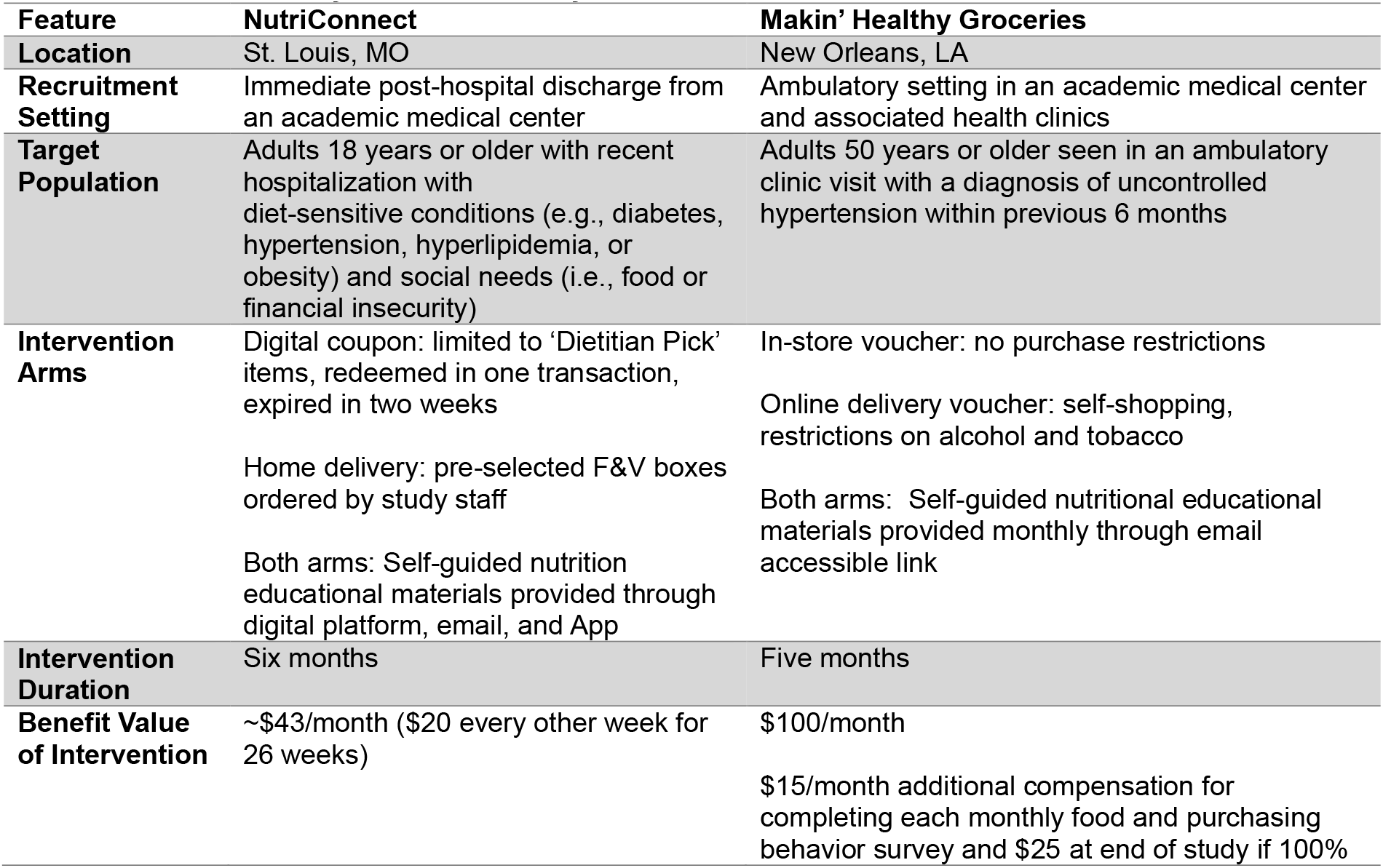
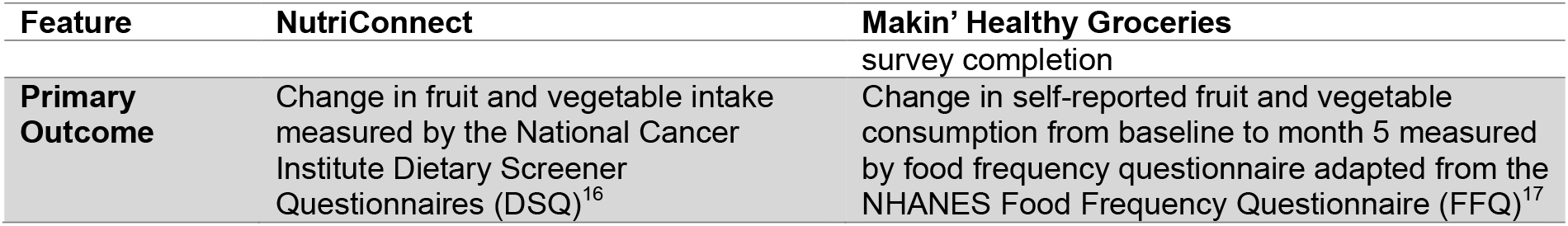
Cross-Site Study Feature Summary.

### Intervention Arms and Delivery Modalities

Each trial implemented multiple study arms to compare different methods of food benefit delivery, behavioral engagement, and purchase control.

- NutriConnect tested three arms:
  1. *Digital Coupon*: Participants received a $20 digital Schnucks coupon every other week for 6 months. This coupon was delivered via email, and its use was restricted for produce and healthy groceries labeled as “Dietitian Pick” at Schnucks. The Dietitian Pick program is an existing system within the grocery chain with items selected by a dietitian employed by the grocery chain. Coupons expired in two weeks and had to be used in a single transaction.
  2. *Home Delivery*: Participants received a fruits and vegetables (F&V) box (fresh, canned, and frozen), equivalent in value to $20, delivered to their home every other week. The home delivery was set for 6 months. Food choices were guided by registered dietitians and informed by patient-centered input. There were six box choices rotating through the intervention phase. The box purchase was made by the study team through an online platform for home delivery.
  3. *Usual Care (Control):* Participants received standard resource referrals to available community services without specific food benefits.

Participants in both intervention arms also received self-guided educational materials via email and were encouraged to engage with Schnucks’ loyalty program and App tracking dashboard. These educational materials were developed through a previous similar project and modified for NutriConnect.

- Makin’ Healthy Groceries tested two arms:
  1. *Brick-and-mortar voucher*: Participants received $100/month debit card style voucher for 5 months to be redeemed in person at grocery partner locations. Restrictions on in-store purchases were not possible with the in-store voucher. The voucher was not required to be used in one transaction, but funds did not roll over to the next month.
  2. *Online voucher delivery*: Participants received $100/month digital credit for 5 months through an online grocery shopping and delivery platform. Orders were delivered to their homes. Contrary to the in-store arm, purchases were restricted to food, beverage, personal care, and household items. Participants were required to navigate a web-based system, share payment details (for service fees, optional delivery tips, or additional purchase costs above the voucher amount), and accept potential substitutions. The credit was not required to be used in one transaction, but funds did not roll over to the next month.

Participants in both arms received educational resources regarding healthy food purchasing and health impacts. . Materials were co-developed prior to the study with patient advisory groups to enhance accessibility and relevancy and were accessible on a webpage. Participants received an email each month containing a link to the resources.

### Operational Features and Value Alignment

Across both studies, teams sought to ensure comparability in benefit value across intervention arms while allowing variation in access modalities and behavioral incentives to enable meaningful comparative evaluation. Implementing the intervention arms required upfront technical planning to establish the systems needed for participants to access study funds. Throughout, there was a deliberate emphasis on integrating with existing food retail systems rather than building parallel or novel distribution channels.

## RESULTS

Notable findings for each dimension of the RE-AIM framework are presented. This approach highlights contextual factors influencing real-world implementation. Particular attention is given to how key program design elements—type of voucher system, the presence or absence of purchasing restrictions, and flexibility to make multiple purchases throughout the month—affected implementation fidelity, participant experience, and overall sustainability.

### Reach

NutriConnect:

- Reach was measured by the proportion of any respondents’ opening at least one push/email communication.
  - The use of existing retail infrastructure (e.g., Schnucks Rewards Program) enabled broader potential reach, but digital enrollment and mobile App setup steps posed access barriers.
  - Digital barriers limited reach. Participants struggled with locating emails, “clipping” coupons, and locating Dietitian Pick items.

Makin’ Healthy Groceries:

- Reach was measured by 1) frequency and amount of voucher use each month, 2) engagement with educational materials, derived from frequency of interactions with the educational website.
  - Participants’ pre-existing familiarity with the grocery store may have reduced barriers to engagement and improved reach in the in-person shopping arm compared with the online arm.
  - Educational materials were co-designed with a patient advisory group, increasing cultural and linguistic relevance.
  - Quantitative indicators (e.g., voucher use, web click data) suggest interest in educational content related to cooking and shopping but less frequent engagement with instructional materials on using the voucher system—particularly for the online group.
  - Adults in both groups encountered technological difficulties due to user interface barriers and/or reduced digital literacy, reducing reach and engagement.

### Effectiveness

NutriConnect:

- Effectiveness was measured by the between-group differences in changes in F&V intake (primary outcome) between baseline and 6-month follow-up, which was assessed through a FFQ administered by phone. A key secondary outcome, food security, was also assessed by phone interviews. Results regarding intervention effectiveness will be described elsewhere.

Makin’ Healthy Groceries:

- Effectiveness was measured by the between-group difference in changes in self-reported food consumption for different food groups including fruits, vegetables, dairy, meat and seafood, desserts and sweets, via online monthly FFQ^17^ between baseline and 5-month follow up, as well as participant reported feedback. Lack of comparable data between in-store and online platforms hindered full program comparative assessment that would have allowed the study team to verify concordance between purchase patterns and consumption behaviors.

### Adoption

NutriConnect:

- Adoption was constrained by:
  - Challenges aligning study intervention specific requirements within Schnucks’ existing categorization system. For example, coupon restriction is only applicable at department level (e.g., produce), which would have prevented participants from using the coupon for canned and frozen fruits and vegetables. This challenge was addressed by limiting the coupon to “Dietician Pic” items - an existing Schnucks program).
  - Challenges related to Schnucks staff effort (for example, the Schnucks communication and marketing team had to set up specific coupon scheduling for the study).
  - Early lessons led to iterative technology adjustments to improve the coupon group’s user experience. For example, providing a coupon schedule to study participants informed them when to expect coupons).
- Adoption was supported by: Strong institutional support from Schnucks.

Makin’ Healthy Groceries:

- Adoption was constrained by technological and operational challenges:
  - Store clerks lacked training to process vouchers efficiently, leading to an unstandardized participant shopping experience.
  - Some online participants were hesitant to share personal financial data as standard protocol required to establish an online shopping account.
  - Data logistic systems were not mature for real-time troubleshooting, leading to delays in transaction verification.
- Adoption was supported by positive participant feedback such as:
  - *“I would like to thank you for the opportunity to participate in this program. It has enabled my purchasing power for groceries to increase over the length of this program*.*”*
  - Adoption of the brick-and-mortar arm was supported by participants who mentioned familiarity with the grocery chain locations and its operations.

### Implementation

NutriConnect:

- Implementation metrics were measured by weekly coupon redemption trends, engagement with coupon text reminders, coupon retrieval, and home delivery engagement.
  - Ongoing adaptations (e.g., coupon delivery text push reminders, simplified coupon retrieval process) improved coupon usage during the study period.
  - Coupons were tied with participants’ store Award account (phone number, email, etc.). This affected engagement when participants were, for example, unable to find the coupon email, having difficulty in “clipping” the coupon to redeem, or failing to redeem before coupon expiration. These factors frustrated participants, as identified through bi-weekly check-ins with participants.
  - Cost was assessed by capturing time devoted to implementation (i.e., not research-related activities) for both the study team and the partners (e.g., Schnucks team).

Makin’ Healthy Groceries:

- Implementation assessment was based on observations made by the research team and feedback provided by participants at different phases spanning patient enrollment, engagement and intervention.
  - Enrollment platform issues (e.g., user interface/user experience, mobile compatibility, cumbersome login processes) required rapid platform customization, delaying recruitment.
  - Manual tasks to support the technical infrastructure (e.g., assigning IDs, external communication systems, non-integrated randomization, lack of customization) led to a high staff burden.
  - Participants in the online group raised concerns over technical assistance and platform unfamiliarity.
  - Stores could not implement voucher controls or purchasing restrictions to block ineligible purchases (e.g., alcohol and tobacco). The feedback loop to identify ineligible purchases was not adequate due to ongoing technology improvements.

### Maintenance

NutriConnect:

- Technical enhancements for NutriConnect (e.g., automated coupon delivery, reminder systems) demonstrated potential for durable system integration.
- Restriction of food purchase to ‘healthy’ foods was tied to existing initiative (Dietitian Pick items).
- Ongoing cost evaluations will inform broader sustainment and scalability planning.

Makin’ Healthy Groceries:

- Program sustainability relies on continual funding for vouchers, staff, and technology supported systems.
- Dual methods of implementation (online vs. in-store) increased the cost for maintenance but also allowed greater flexibility to meet participants’ needs.

Table 2, below, outlines and expands on various findings from the comparative analysis.

**Table 2.**
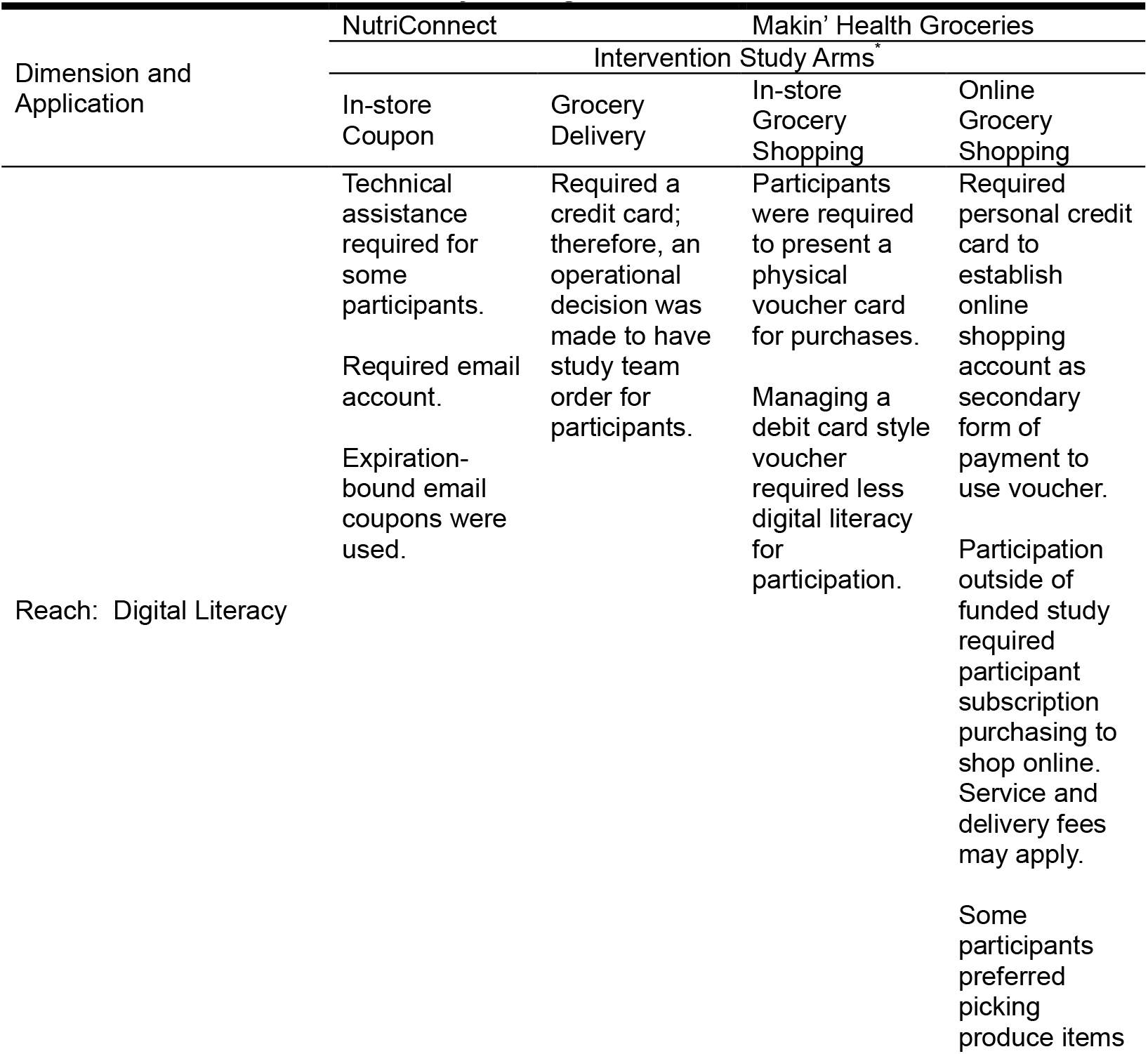
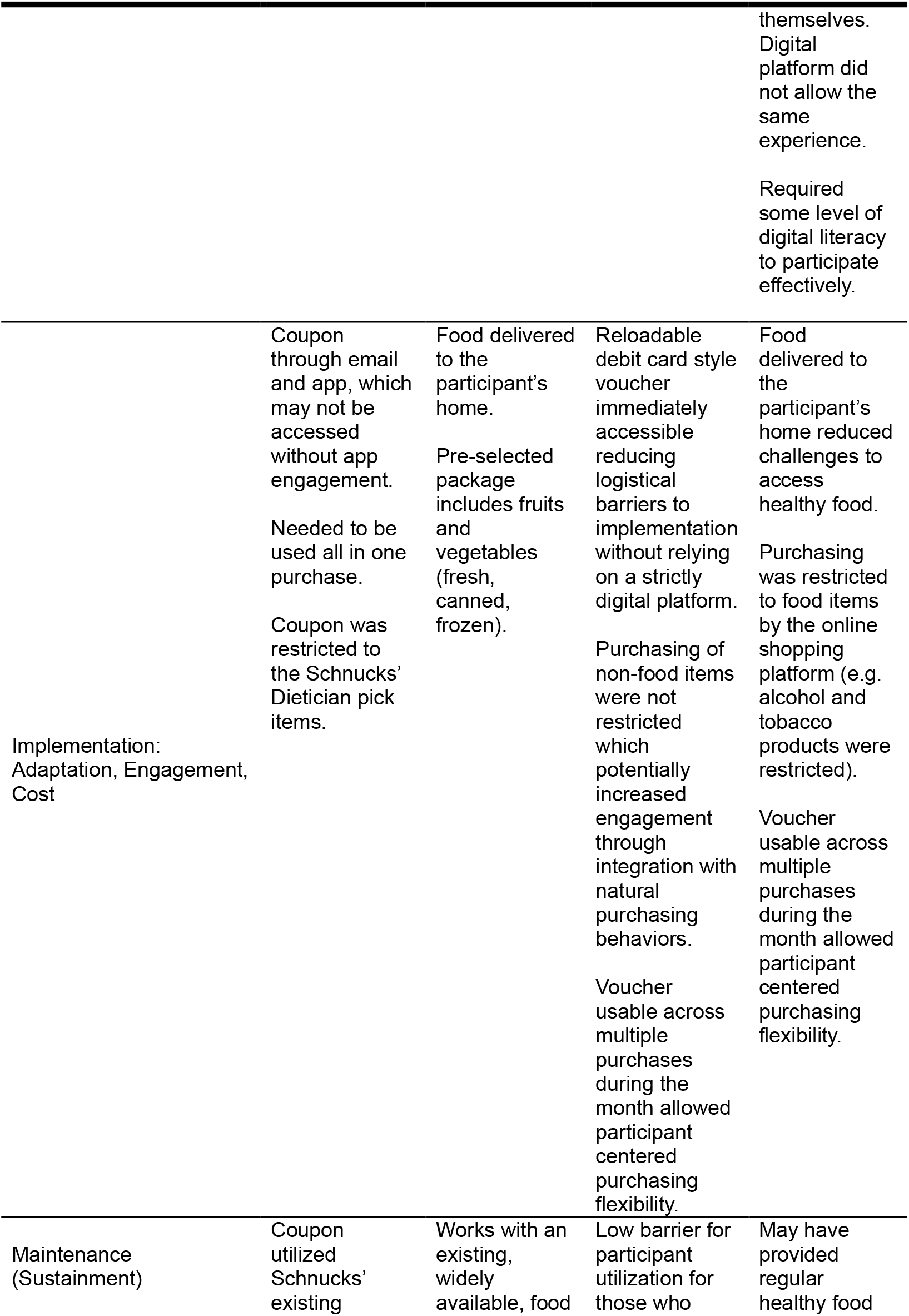
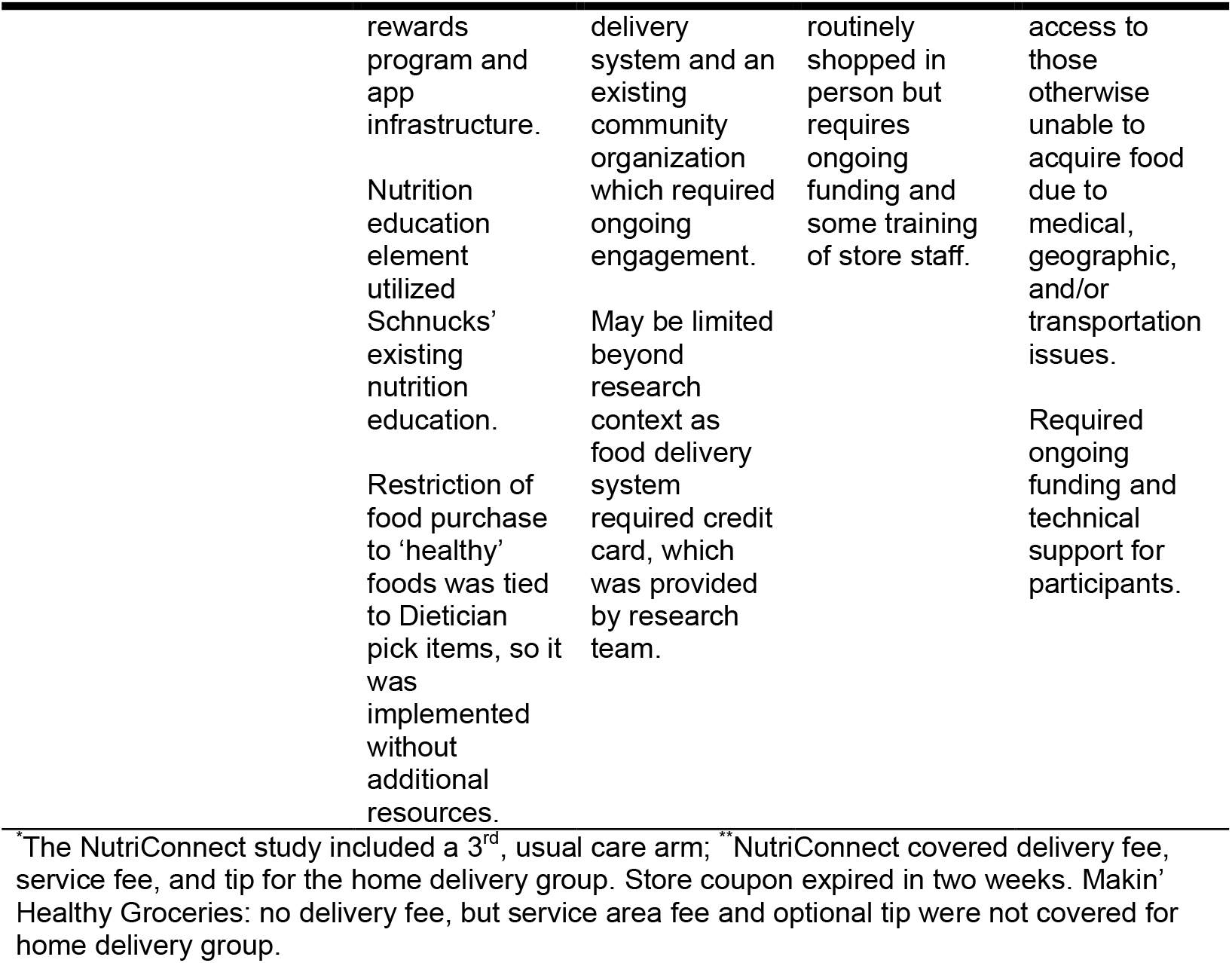
Select Notable Cross-Study Findings.

## DISCUSSION

This comparative analysis of two pilot interventions—*NutriConnect* and *Makin’ Healthy Groceries* — offers a window into the heterogeneity of FIM program design and delivery, in the context of FIM models embedded in diverse community and healthcare ecosystems. While each pilot was contextually specific, their differences— including target populations, implementation strategies, and infrastructure—revealed several insights into conditions required for successful implementation, fidelity, and long-term sustainability of FIM interventions.

Applying the RE-AIM framework enabled a structured comparison of key implementation domains and illuminated both cross-study lessons and persistent challenges in the field. These findings suggest that institutionalization of FIM programs within healthcare and retail systems will require not only strong partnerships with the food retail market, healthcare, and communities but also shared investments in technology and training to reduce friction and enhance operational efficiency.

### Reach: Digital and Structural Barriers Shape Participation

A core insight from both interventions was that the implementation modality significantly affected trial reach.

For example, in the *Makin’ Healthy Groceries* trial, participants demonstrated strong engagement with the brick-and-mortar shopping experience, citing familiarity and autonomy as key reasons. In contrast, lower initial engagement was found in the online arm due to digital platform navigation challenges, but engagement increased once familiarity was established. The *NutriConnect* trial faced similar challenges. Despite integrating benefits with the existing Schnucks loyalty platform, reach was limited by the requirement for an active email account, some participants’ low familiarity with the Schnucks loyalty and reward program, and low willingness to complete all purchases in a single transaction which was a requirement of the study These findings underscore the importance of aligning intervention delivery models with the lived experiences, natural purchasing behavior, and technological capabilities of the target population. Digital tools should be designed to be intuitive, culturally relevant, and well-supported by human navigation assistance. Tailoring delivery mechanisms to existing behaviors (e.g., in-person shopping) and integrating human support may improve engagement. This further highlights issues related to the target population, where patients in outpatient care may be more able to engage with interventions than those recently discharged from acute care.

### Adoption: System Readiness and Operational Integration

Successful adoption of the interventions varied across study arms and was highly dependent on the readiness (e.g., technical infrastructure) of grocery retail and digital partners. Adoption of *Makin’ Healthy Groceries’* in-store intervention was hindered by operational gaps such as limited store clerk training, lack of automated transaction tracking, and an inability to restrict the purchase of ineligible non-food items. These challenges point to the need for early investment in operational alignment, workflow integration, and staff engagement strategies within retail environments. Even well-intentioned partners may lack the systems or training protocols to fully support research-oriented programs. *NutriConnect’s* adoption, in contrast, benefited from tighter integration with Schnucks’ data systems, including linked coupon redemption and product curation. However, enrollment still required participants to complete several digital steps which limited adoption and implementation. Both studies highlighted that staff engagement, system compatibility, and real-time support were critical for optimizing implementation fidelity.

### Implementation: Flexibility and Problem-Solving Are Essential

Both studies encountered substantial implementation challenges related to their respective digital infrastructures. In the *Makin’ Healthy Groceries* trial, the enrollment and engagement platform, which was originally designed for healthcare data integration, required extensive customization to support participant facing protocols, response tracking, survey administration, and communication. Participant feedback pointed to practical concerns such as font size and password recovery that may have disproportionately affected underserved populations such as those with reduced digital literacy. The *NutriConnect* intervention arms similarly required the development of a separate communication and coupon distribution module for the trial, ongoing modifications to improve coupon distribution, and redemption tracking.

Both pilots demonstrated strong capacity for real-time adaptation. User experience issues were rapidly identified and resolved through logistical improvements, manual workarounds, and participant feedback loops. These adaptations highlight the critical importance of flexible technology platforms, robust technical support, and a willingness to iterate during implementation. *NutriConnect’s* adaptions provided strategies to optimize coupon retrieval and communication processes in collaboration with Schnucks and community partners, which are now being scaled into additional programs.

Taken together, these learnings reinforce the need for proactive usability testing and human-centered design approaches in FIM program development particularly when interventions rely on digital platforms to deliver services or track behavior.

### Maintenance: Financial, Technological, and Institutional Sustainment

Maintenance emerged as a critical challenge for both trials. The costs of sustaining FIM intervention distribution represent a significant ongoing investment, not inclusive of staff salaries, technology maintenance, and research infrastructure. Technology platforms, while critical to data management and engagement, also require long-term support and funded staff. Improvements made during the *NutriConnect* trial, such as enhanced coupon management and participant tracking systems, have since been adopted in other programs (e.g., a local AHA chapter initiative has adopted the coupon system originally established through the *NutriConnect* implementation)—indicating potential for scalability. These technical enhancements are now being used in other food assistance or FIM programs. However, the experiences from both studies suggest that widespread maintenance and replication of FIM programs will likely depend on integration into existing institutional, retail, and healthcare workflows, alignment with reimbursement models, and continued investment in digital infrastructure.

Early planning for maintenance must include cost modeling, reimbursement pathways, and opportunities for integration into existing platforms such as electronic health records, loyalty apps or rewards programs, and community health worker workflows. These findings highlight the urgent need for relevant policies that support payment structures (e.g., Medicaid waivers, Center for Medicare and Medicaid Services investment) and cross-sector partnerships that can absorb costs and sustain delivery over time.

### Cross-Cutting Lessons and Future Directions

This comparative analysis reveals not only the heterogeneity of FIM program delivery but also consistent themes of implementation in context:

1. *Technology can be a double-edged sword*: While digital platforms allow for control and automation, they can create barriers for some vulnerable populations. Hybrid models that pair digital technology with human support likely offer the best balance.
2. *Retail partnerships are essential but complex*: While grocery chains offer scale and reach, meaningful adoption requires strong data and logistical integration, staff training, and clear communication protocols.
3. *Participant experience is as important as the outcome*: Trial designs should emphasize autonomy, trust, and familiarity to drive engagement and behavior change more effectively than overly prescriptive models.
4. *Maintenance and sustainability require more than cost savings*: Even with promising outcomes, programs need policy alignment, institutional champions, and robust data infrastructure to become permanent offerings.

As the FIM landscape continues to evolve from pilots to larger scale trials to policy implementation, these insights provide a roadmap for program design, partnership models, and equitable implementation that target the most at risk community members.^11,13,18,19^ Future research should explore: 1) how adaptive intervention designs and mechanisms can integrate FIM models into retail systems and simultaneously into healthcare delivery and social service systems, 2) how to develop responsive digital tools for diverse populations, and 3) long-term clinical and economic effects of FIM strategies on health outcomes and healthcare utilization patterns.

## Data Availability

Data will be available upon request

## Non-standard Abbreviations and Acronyms

AHA: American Heart Association
DSQ: Dietary Screener Questionnaire
FFQ: Food Frequency Questionnaire
F&V: Fruits and Vegetables
FIM: Food is Medicine
LPHI: Louisiana Public Health Institute
NHANES: National Health and Nutrition Examination Survey
RE-AIM: Reach, Effectiveness, Adoption, Implementation, Maintenance framework
UMCNO: University Medical Center New Orleans
WashU: Washington University in St. Louis

## Acknowledgements

Duke Clinical Research Institute: Prime institution which led the study with Louisiana Public Health Institute direction by providing administrative and project support as well as statistical analysis and clinical guidance. University Medical Center New Orleans: Led clinical site recruitment and enrollment of eligible participants for Louisiana Public Health Institute’s study. Grocery partners: Provided support for the retail integration of the intervention and purchase data acquisition.

